# Significantly Improved COVID-19 Outcomes in Countries with Higher BCG Vaccination Coverage: A Multivariable Analysis

**DOI:** 10.1101/2020.04.23.20077123

**Authors:** Danielle Klinger, Ido Blass, Nadav Rappoport, Michal Linial

## Abstract

COVID-19 has spread to 210 countries within 3 months. We tested the hypothesis that the vaccination with BCG correlates with a better outcome for COVID-19 patients. Our analysis covers 55 countries, complying with predetermined thresholds on population size and deaths per million (DPM). We found a strong negative correlation between the years of BCG administration and a lower DPM along with the pandemic progression in time. The results from multivariable regression tests with 22 economical, demographic, and health-related quantitative properties for each country substantiate the dominant contribution of BCG administration years to the COVID-19 outcomes. Analyzing countries according to an age-group partition reveals that the strongest correlation is attributed to the coverage in BCG vaccination of the young population and mostly to recent years immunization. We propose that BCG immunization coverage, especially among the most recently vaccinated contributes to attenuation of the spread and severity of the COVID-19 pandemic.

**One Sentence Summary:** BCG vaccination regimes and COVID-19 outcomes

## INTRODUCTION

COVID-19 spread within 3 months to over 210 countries across the globe. The progression of COVID-19 pandemic in each country is estimated by the daily report compiled by the World Health Organization (WHO) and publicly available through the Worldometers online resource (*1*). The country-specific reports provide statistical information on the number of tests performed, the COVID-19 confirmed cases, the number of deaths, and the cumulative state of patients hospitalized in serious and critical conditions. Along with the spread of the pandemic, most countries imposed a policy of social distancing and other measures to mitigate COVID-19 (*2, 3*). Despite the intense effort, the knowledge on the COVID-19 SARS-CoV-2 virus is fragmented and key epidemiological parameters are still missing (*4-9*). With over 250k reported deaths and an average number of 33 deaths per million (DPM, as of May 5th, 2020), the death toll remains a reliable measure for monitoring the spread and progression of the disease across countries (*10*). While for some European countries such as Belgium, Spain, Italy, and the UK the DPM is >400, other infected countries (e.g., Brazil, Turkey, Israel, Hungary) are closer to the world average. The large differences in COVID-19 outcomes, even among neighboring countries (e.g., Spain and Portugal) are not likely to solely reflect differences in the regulations imposed by each country at the initial phase of the pandemic (*11, 12*).

In this study, we tested the possibility that the extent and spreading of COVID-19 cases are associated with the status of tuberculosis (TB) immunization across the world. The Bacille Calmette-Guérin vaccine (BCG) contains a live attenuated strain of *Mycobacterium bovis*, is widely used to eradicate TB and was among the most broadly used vaccination throughout the 20th century in neonatal and young children (*13, 14*). Despite its wide use in many developing countries, about 1% of the world population is infected with TB each year (*15, 16*). While a constant reduction in death due to TB is observed over the last decade, the annual deaths exceed 1.5M (*17*), with a substantial overlap with people that are HIV infected (*18,19*). A high prevalence of TB in some countries was attributed to an increase in diabetes mellitus (*20*), and fast changes in population composition (*21*). Currently, the BCG vaccine is provided to the entire population in most countries with high TB incidence (*22*). Over the last two decades, numerous countries changed their policy and restricted BCG immunization policy to non-native born migrants from high TB burden countries (*23*). Notably, numerous epidemiological and immunological studies demonstrate that BCG vaccination results in reduced morbidity and mortality to subsequent infections, presumably by an effect on the immune response (*24*). In this study, we questioned whether the long-range impact of the BCG vaccination regimes and the varied fraction of the population that is covered by BCG in different countries are linked with an improved COVID-19 outcome.

## RESULTS

### Unified COVID-19 pandemic across countries by alignment key dates

Analyzing the trends of the COVID-19 pandemic across different countries is prone to variability on the actual date in which countries were exposed and where COVID-19 fatality is recognized and reported. To increase the robustness of our analyses, we focused on countries with a minimum population size of >3M people and mandatory 3 DPM at the analysis date (17 April 2020). Altogether, 55 out of 210 countries covering 62.9% of the world population complied with these thresholds. A regional partition of these countries is shown in **Fig. 1**. For detailed information on the countries included in the analyses, see Supplemental **Table S1**.

**Fig. 1.**
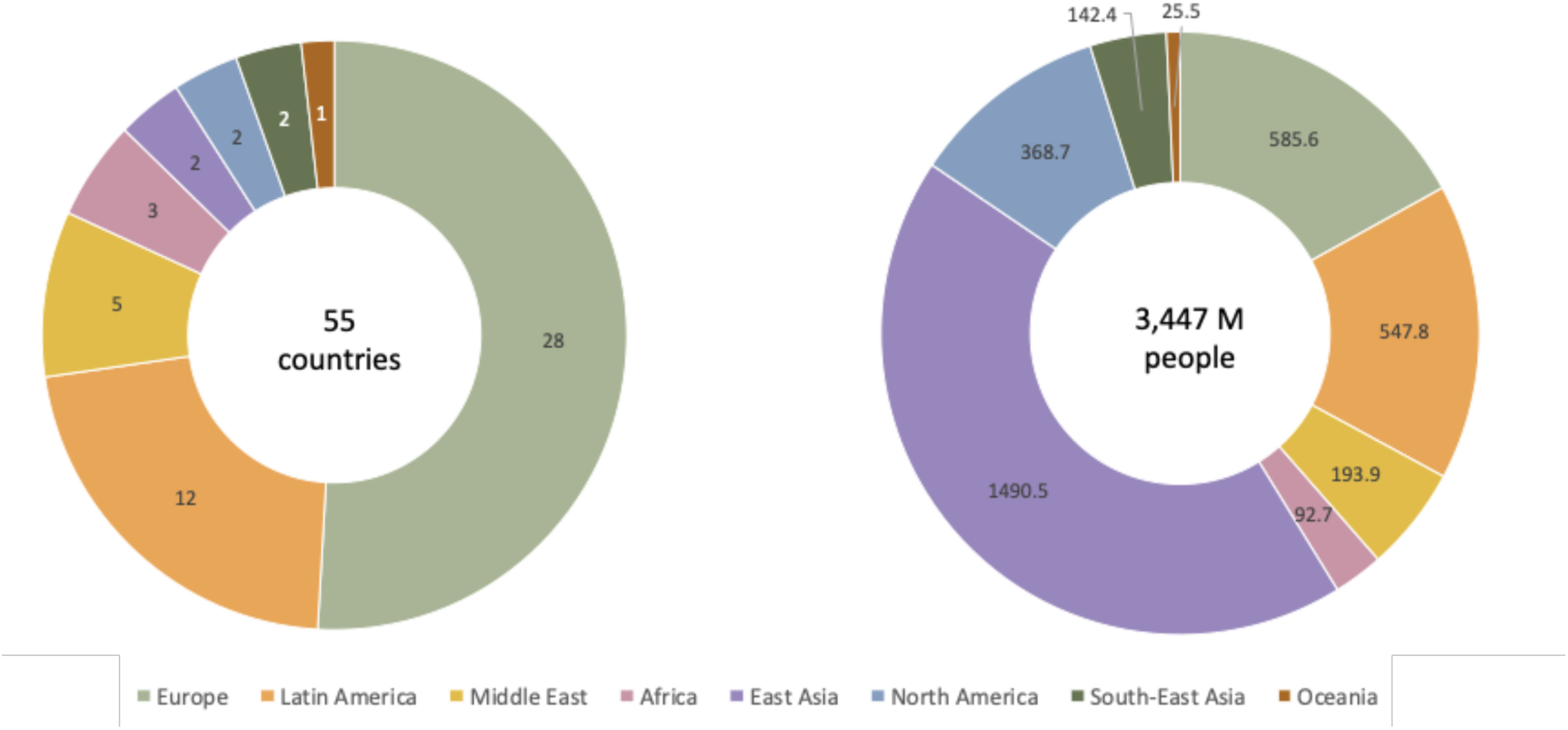
Countries analyzed in this study by geographical regions. Countries that comply with the predetermined threshold for the population size and a minimum DPM at the analysis date are included. These countries (55 total) are partitioned by their geographical regions (left) and the cumulative population size within each region (right).

### BCG administration years are negatively correlated with COVID-19 outcomes

First, we analyze COVID-19 outcomes as the difference in death per millions (DPM) or cases per million (CPM). **Figure 2** shows strong and significant correlations between COVID-19 outcomes and the number of years of BCG administration. The analysis is performed 20 days following 2 different alignment key dates (defined by DPM = 0.5 and DPM = 1.5). We observed a strong negative correlation with both DPM and CPM as a function of years of BCG administration. For DPM outcome, we observed a correlation of R = −0.48 (p-value = 0.00083) when aligned at DPM >= 0.5 (**Fig. 2a**) and R = −0.47 (p-value = 0.001) when aligned at DPM >= 1.5 (**Fig. 2b**). Similarly, considering the CPM rather than the DMP results is a similar trend with slightly lower correlation and statistical strength. Specifically, at 20 days post alignment, we observed a correlation of R = −0.37 (p-value = 0.012) when aligned at DPM >= 0.5 (**Fig. 2c**) and R = −0.34 (p-value = 0.02) when aligned at DPM >= 1.5 (Fig. 2d). Repeating the analysis at various time points along with the progression of the disease (e.g., 30 days after the aligned key date) resulted in a similar trend of highly significant negative correlation between COVID-19 outcomes and years of BCG administration (**Fig. 3**). Note that at 20 days post alignment of DPM >= 0.5, as many as 45 out of 55 of the countries provided information. The rest of the countries were at an earlier phase of the pandemic.

**Fig. 2.**
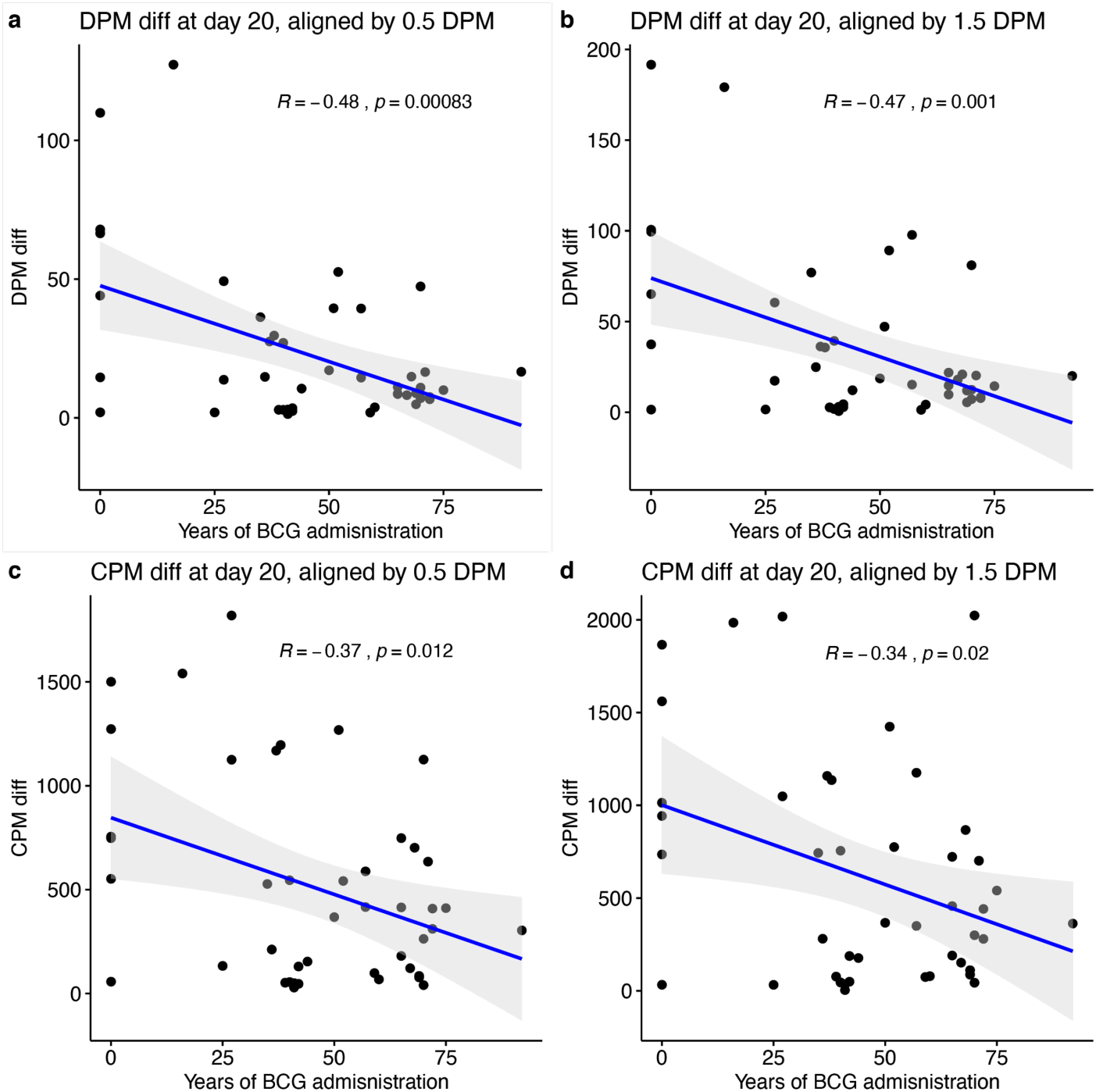
Statistical analysis of the countries COVID-19 outcomes and years of BCG administration. All correlations were measured at 20 days following the alignment key date. Correlations of years of BCG administration with DPM = 0.5 **(a)** and DPM = 1.5 **(b)**. Correlation with CPM diff. at 20 days when the key date was defined as CPM = 0.5 **(c)** and CPM = 1.5 **(d)**. DPM diff. and CPM diff. are calculated by the differences in the numbers from the measured date to alignment date. Shaded areas represent the 95% confidence intervals.

To test the generality of our observations we repeated the analysis at a broad range of time points along with the progress of the disease in the different countries, starting from the 10th-day post the alignment key date and showing the trends in 5 days intervals (10 to 40 days post alignment, **Fig. 3**). For this analysis, we tested the outcome of COVID-19 confirmed serious/critical cases (SPM) and the number of recovered (RPM), in addition to the DPM and CPM. The results of the DPM and SPM show a highly significant association at all time points, corroborated by the robust results obtained from permutation tests (**Table S2**).

**Fig. 3.**
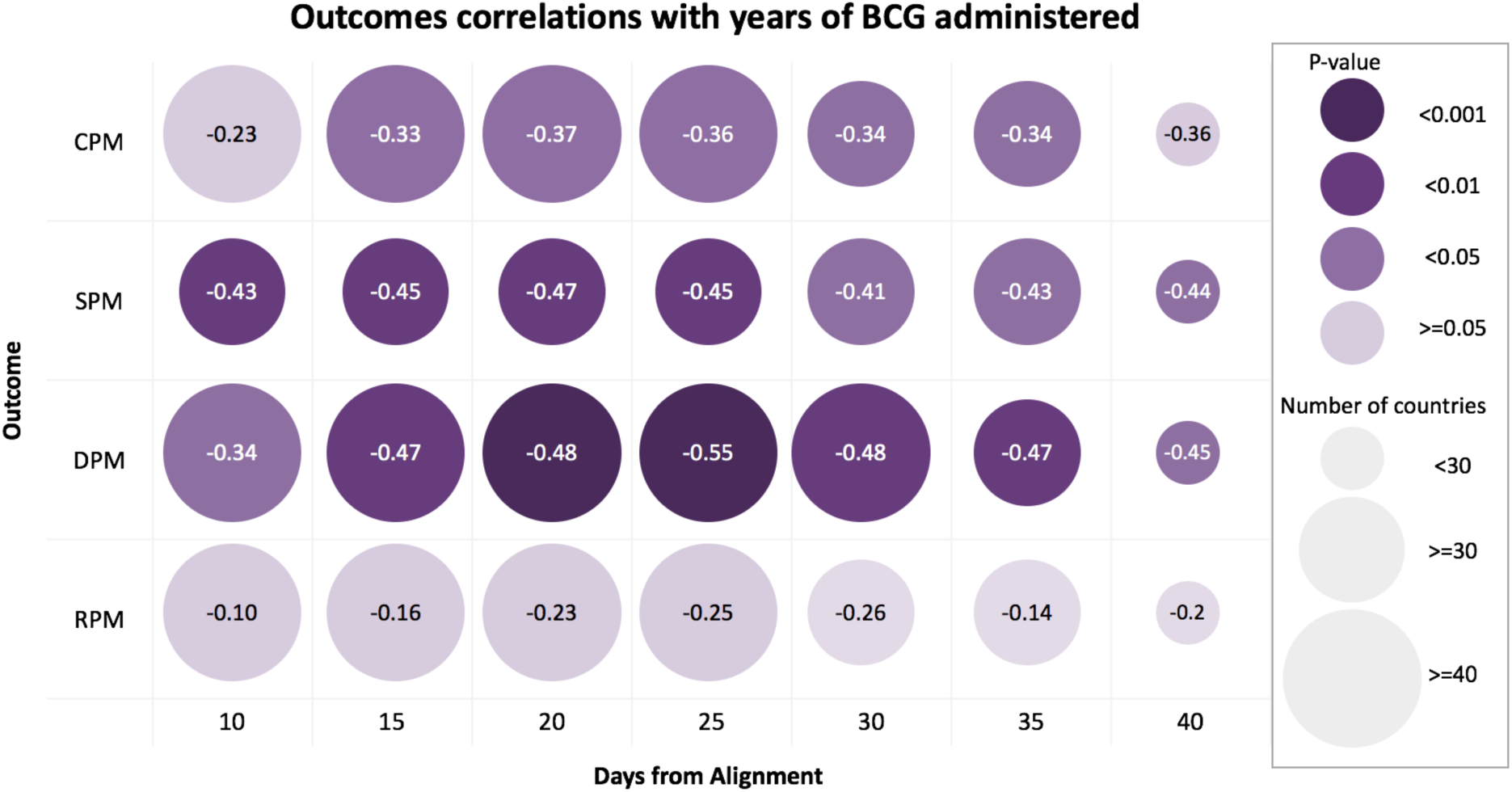
Outcomes correlations with years of BCG administered. Each row represents a different COVID-19 outcome. Each column represents the time interval of the outcome from the alignment date. Circle size depicts the number of countries, and color represents the statistical significance of the correlation with darker purple color indicating a higher significance. Values in circles are the correlation estimates. Note that all correlations are negative.

### Multivariable analysis reveals the strong contribution of BCG administration to the statistical signal of COVID-19 outcome

Countries differ in many quantitative measurements like population size, Gross Domestic Product (GDP), lifespan, median age, and more. To control for some of the potential confounding factors, we included numerous demographic values for a multivariable linear regression. The analysis included 22 demographic, economical, and health-related country-based variables. The results show that the number of BCG administration years ranks consistently within the top two most significant coefficients and is within the top coefficients with the larger effect (as measured by the normalized beta coefficient, out of 22 coefficients) (**Fig. 4**). The results are consistent among the different times observed: 10 days-ranks 5th largest, p-value = 0.0028, beta = −0.67; 20 days-ranks 4th larger effect, p-value = 0.00198, beta = −0.814; 30 days-ranks 7th largest effect, p-value = 0.00698, beta = −0.724. Unsurprisingly, a strong positive beta coefficient is associated with the median age. Cancer percentage is also significant and it is probably a reflection of the lifespan and the tight connection of cancer occurrences and advanced age. The combined contribution of gender, chronic disease prevalence, and economy in the multivariable analysis will not be further discussed.

**Fig. 4.**
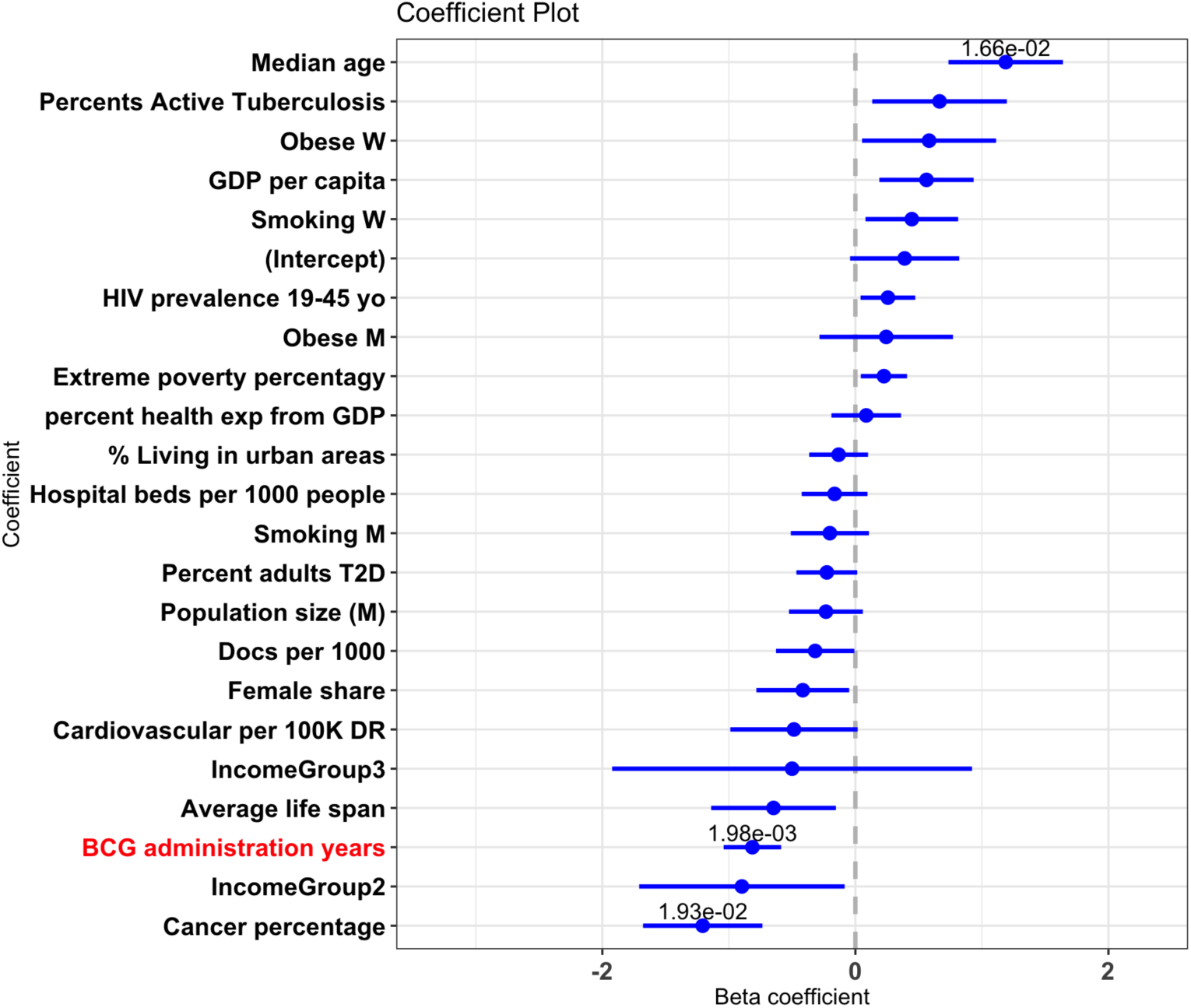
Multivariable analysis of country-centric quantitative data. Beta coefficients of the normalized multivariable linear regression for DPM at day 20 are shown. Blue lines represent the coefficients’ 95% confidence intervals. P-values were added for all variables with p-value < 0.1.

### Highest correlation with BCG age coverage applies to the most recently vaccinated

We next investigated the relevance of age groups to the observation showing that years of BCG administration are strongly correlated with better COVID-19 outcomes (**Fig. 2 & 3**). As it is evident that countries differ drastically in their median age, we inspected the age partition of the population for each country. For instance, while in Ecuador the median age is 27.9, it is 47.3 years in Italy. **Fig. 5** shows the correlation of total years of BCG administration with DPM difference, within the given timeframe of the age group according to the country-based age composition, 20 days post alignment key dates. The population in each country was partitioned to young (<24 years of age), working-class (25-64 years) and old (>65 years). The correlation with the young age group (testing at 20 days) shows the highest significance with R = −0.55, p-value = 9E-05 (**Fig 5a**). The highly significant negative trend repeats among all dates tested within the 10-40 days post alignment. The correlation with the age group of 25-64 years (testing at 20 days) is also significant with R = −0.31 with a weaker significance (p-value = 0.039) and remains significant throughout the 40th-day post alignment (**Fig 5b**). Remarkably, for the old age group (>65 years) for all the time-frame tested, the correlation is negligible and insignificant (**Fig. 5c**).

**Fig. 5.**
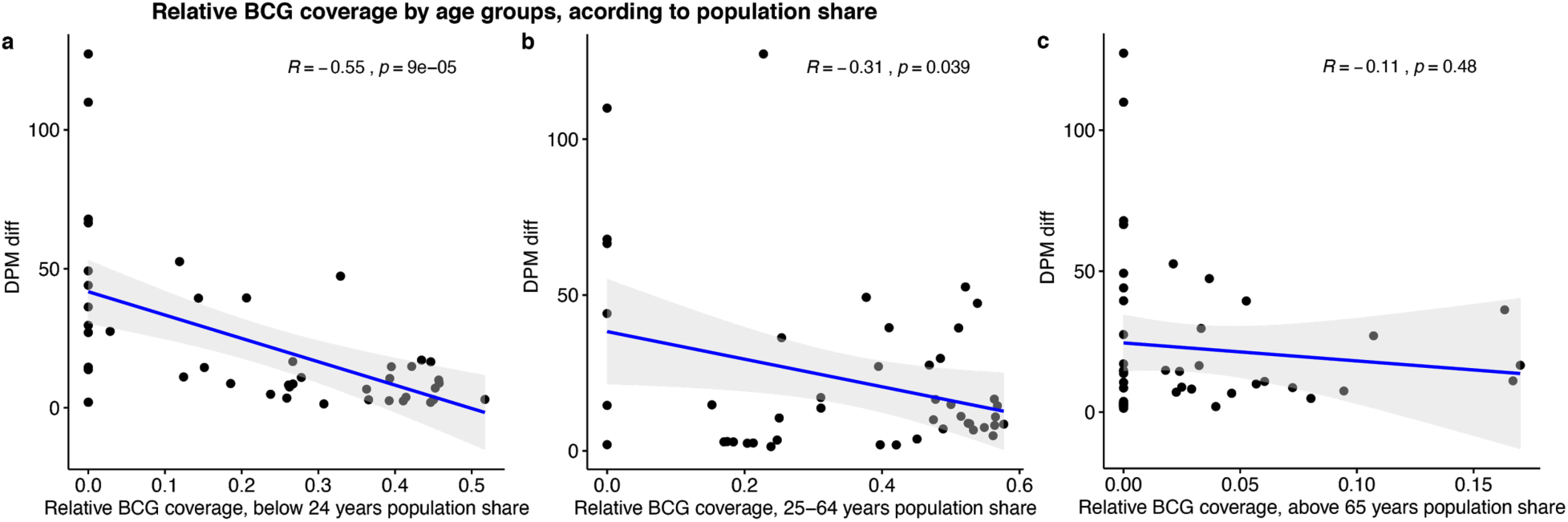
Correlations of relative BCG coverage with DPM difference among varying age groups. All correlations were measured at 20 days following the DPM>= 0.5 alignment key date. Relative BCG coverage within the young age group **(a)**; Relative BCG coverage within the working class age group **(b)**; Relative BCG coverage within the old age group **(c)**.

We further tested the significance of the correlation with different outcomes, by age group according to population share, along different time points post alignment (**Fig. 6**). Comparing the significance by −log10(p-value) clearly shows the contribution of the young age group to the strong negative correlation observed (**Figs 5a & 5c**) for the outcomes at 10-40 days post alignment. Unsurprisingly, the recovered per million (RPM) has a significant negative correlation among the young, as it is a factor of a low number of cases in respective countries. Interestingly though, the recovered per million (RPM) is below significance at day 10 (**Fig. 6a**), which is consistent with the epidemiological survey reporting the long time needed for recovery (*25*). The middle age group (**Fig. 6b**) is mostly insignificant and shows a borderline significance for the DPM as an outcome. All observations regarding the elderly (**Fig. 6c**) are insignificant. We conclude that the elderly group does not contribute to the strong correlation with BCG administration. A minority of outcomes presented a positive correlation, all insignificant (marked by astrocytes in **Fig. 6**).

**Fig. 6.**
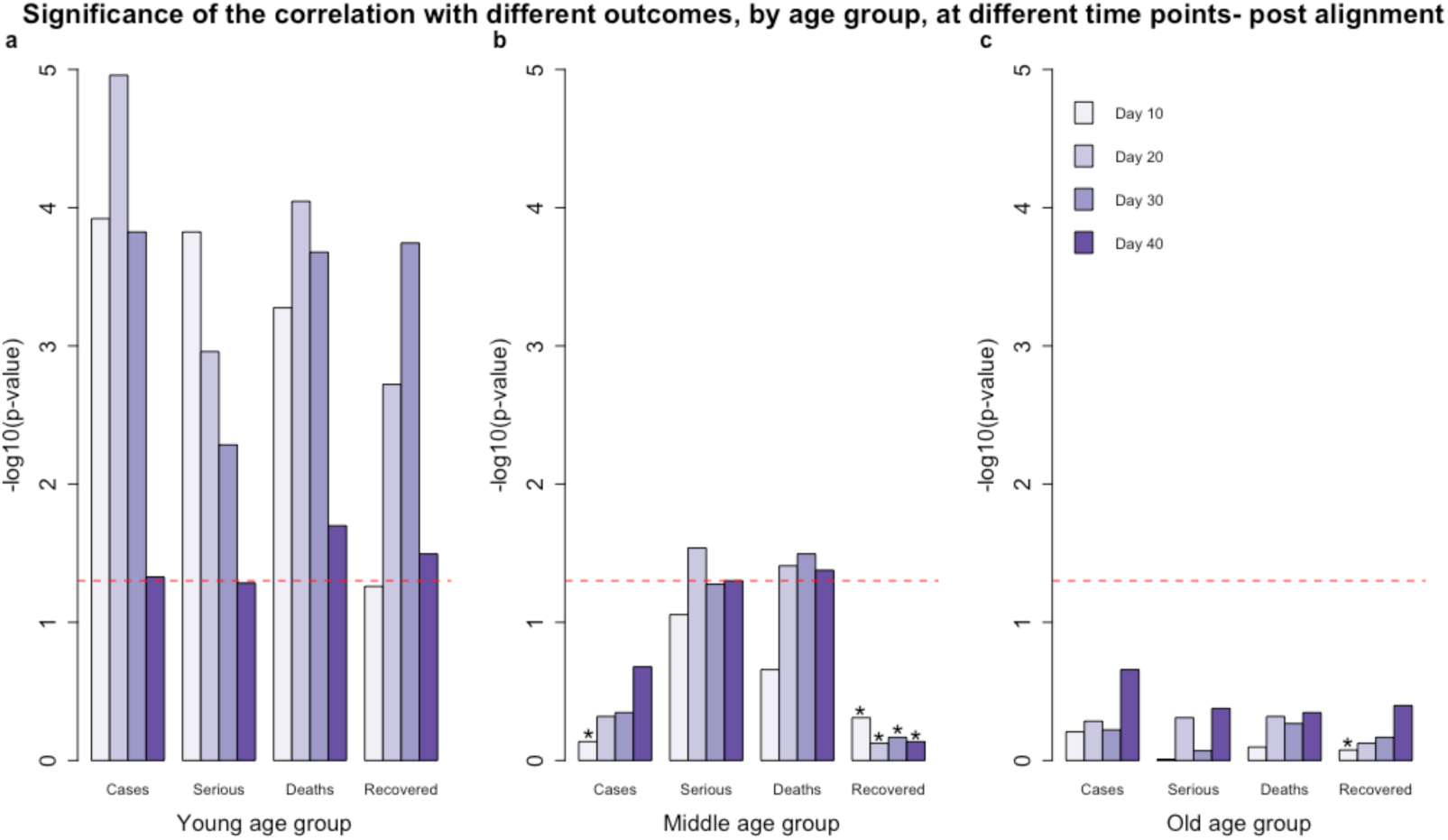
Statistical significance of the correlation of years of BCG administration and COVID-19 outcomes. The histogram shows the statistical significance of the correlation of BCG years of administration and the 4 different COVID-19 outcomes. Days from the key alignment date are colored from light to dark purple (10 to 40 days). The statistical significance is shown as −log10(p-value), the dashed red line indicates p-value of 0.05. The population of each country was divided into three age groups, weighted by population share: young (0-24 years) **(a)**, middle age (25-64) **(b)**, and old age group (>65 years) **(c)**. Astrocytes represent outcomes with a positive correlation.

**Fig. 7.**
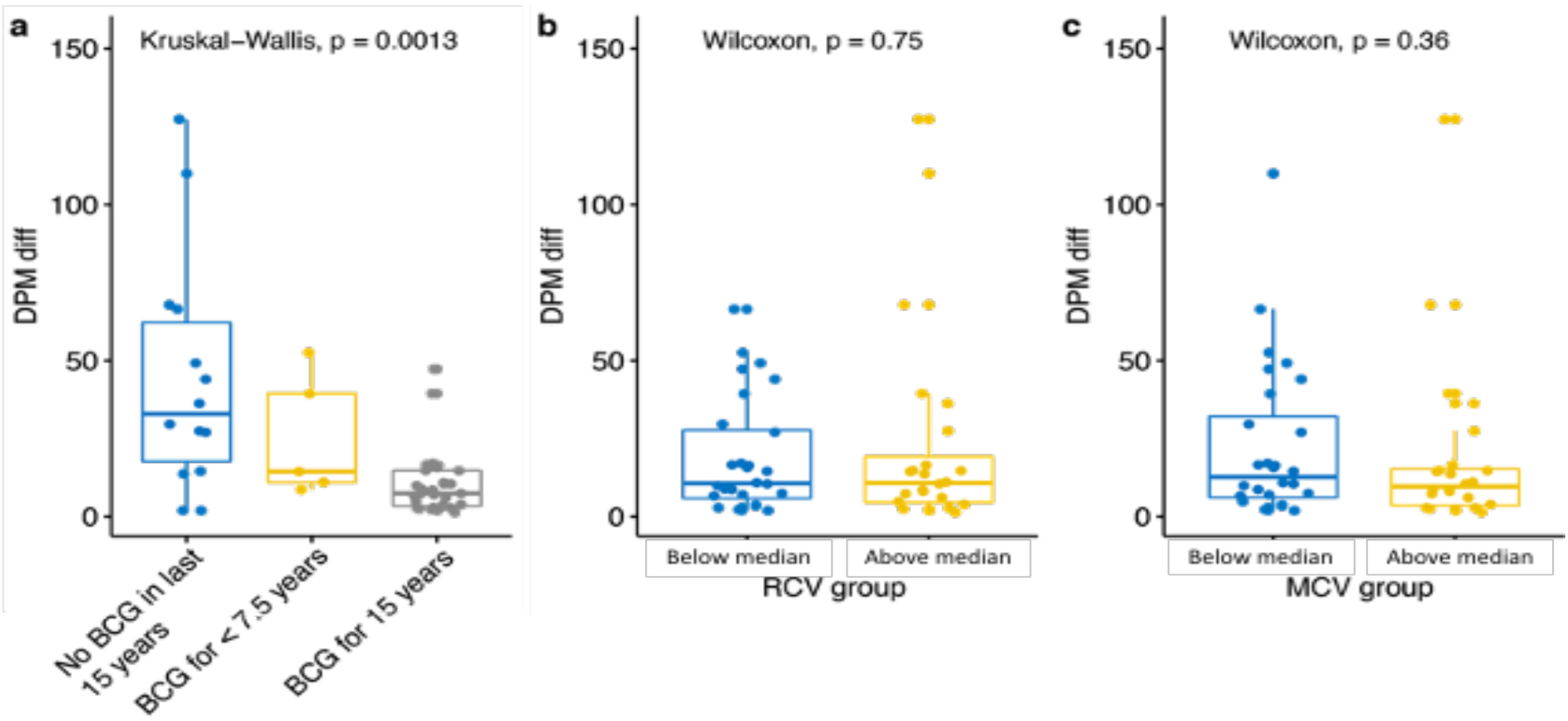
Immunization coverage for 15 recent years. The DPM diff. (difference in DPM from the value at alignment key date) in countries that have (gray), partially have (yellow), or have not (blue) rolled immunization BCG programs over the past 15 years. The statistical significance values are shown for each pair **(a)**. The Statistics of RCV (Rubella) **(b)** and MCV (Measles) vaccines **(c)** and COVID-19 DPM diff. according to the vaccination coverage. The partition of the countries is according to those above (blue) and below (yellw) median % coverage for each of these tested vaccines. Data covering of the past 15 years (2004-2018) was extracted from WHO reports. The statistical significance values are listed.

The pronounced signal in the young age group led us to investigate whether recent immunization may have a positive effect on the outcome, and whether the signal may be due to the proximity in age group of the young to the time of vaccination. The efficacy of BCG against TB is expected to span approximately 15 years (*26*). We therefore divided the countries into 3 disjoined groups representing their vaccination policies over the past 15 years: (i) countries with mandatory immunization policies over the past 15 years; (ii) countries with mandatory immunization policies, which was applied for less than 7.5 years within the past 15 years; (iii) countries with no mandatory immunization policies over the past 15 years **(Fig. 7a)**. Applying the Kruskal-Wallis test to the outcome of DPM, yielded highly significant results at a range of tested dates post alignment (10-40 days), establishing that countries with BCG immunization policies over the past 15 years have a significantly lower rate of DPM with respect with countries in group (iii). **Figure 7a** shows results for 20 days post alignment, with a p-value of 6.3E-04 between the fully treated set of countries (group i) and the non treated (group iii).

The significant result in the young age group led us to investigate whether others globally used immunizations, against measles and rubella-might have a similar effect to that observed for BCG on COVID-19 outcomes (**Fig. 7a**). While the world overall coverage of MCV1 (measles containing vaccine) is high in recent years, with an average of 88% in 2017-as provided by the WHO, the MCV1 coverage from 1990 shows substantial variation (e.g., 73.8% in India, 80.2% in Italy, 85.7% in Algeria, 89.2% in Belgium)(*27*). To this end, we divided the countries into 2 groups representing their vaccination coverage over the past 15 years (as provided by the WHO)-(i) countries with above median coverage; (ii) countries with below median coverage. Applying the Wilcoxon test, yielded insignificant results at all tested dates (10-40 days post alignment).

We provide an interactive website in https://covi.shinyapps.io/COVID19/ as a useful analytical webtool for single variant statistics, correlations, multivariable analyses and more. The user-friendly platform allows users to change parameters along the pandemic progression by selecting any predetermined outcomes as a reference date for the alignment.

## DISCUSSION

The significant strong correlation between the BCG vaccination and better outcomes for COVID-19 is shown across many countries, covering the majority of the world population (62.9%). This finding is based on an unbiased view of all countries that comply with predetermined thresholds on a minimum number of DPM and population size (see Materials and Methods). We anticipate that with the progression of COVID-19 pandemic more countries that were at an early phase of the pandemic at our analysis threshold (17 April 2020) could be included. Importantly, the strong negative correlation between the BCG administration years and DPM was sustained at a range of time-points (**Fig. 3**). For testing the robustness of our DPM correlation, we included other COVID-19 outcomes that reflect other aspects of fighting the pandemic in different countries. We consider the number of people (at a specific date) that were hospitalized and are in serious and critical condition (SPM) not only as a proxy for the pandemic scale, but also an indirect reflection for the health system capacity, and the stage of the pandemic in the subject country. For countries where the spread of the pandemic is high, the ratio of SPM to DPM is high (e.g. Brazil, Russia, Moldova, and Chile) where this ratio is much smaller for countries that already passed the pandemic growth phase (e.g., Canada, Portugal, Netherlands, and Spain). Using the SPM as a measure, we observed that the trend for BCG administration years observed with DPM was similar and remains significant for SPM (**Fig. 3**).

The negative correlation to COVID-19 positive cases (CPM) is weaker relative to DPM. It reflects the varying capacities of different countries to carry out molecular tests (PCR-based) or lung CT pathology criteria for confirming an individual as a confirmed COVID-19 positive (*28*). More importantly, the policy of who is being tested greatly varies among countries (*29*). We observed a no significant correlation for people that were marked as recovered (RPM). We attribute it to the lengthy process of recovery (*30*). Additionally, the non-standardized definition for COVID-19 confirmed recovery (*31*) reduces the reliability of RPM and eventually affects its statistical power.

Our multivariable analysis highlights the strength of combining a broad range of country-based quantitative observation (e.g., economic measures, number of doctors per 1000 people), and population composition including habits (smoking), exposure to infectious diseases (prevalence of TB) and several major comorbidities (e.g. cancer, diabetes). Many of these individual measures are correlated and may reflect confounding factors. For instance, countries that suffer from a high level of TB (*32*) are also marked by high immigration, social inequalities, high drug or alcohol abuse, and HIV infection (*19*). Most importantly, the multivariable analysis validated the importance and statistical significance of BCG immunization years given all other variables, attributing better outcomes to them. As expected, the median age was also extremely significant, with higher values strongly contributing to the worse COVID-19 outcome.

The detailed information on the number of BCG administration years is used as an additional source of information that is unavailable when a binary definition of BCG immunization is considered. Detailed information on the exact years from the first administered year, the health policy in each country, and the actual immunization coverage (provided by the WHO) allowed a thorough inspection of the BCG vaccination by the population structure (**Fig. 5**). We explicitly tested what component of the BCG immunization is most critical for the COVID-19 outcomes. By partitioning the population to young, middle and elderly age groups, we confirmed that the strongest signal towards COVID-19 outcome is associated with the young (<24 years, **Fig. 6a**), and to a minimal signal by the middle age group (25-64 years, **Fig. 6b**). However, the elderly (>65 years) that are at the highest risk for COVID-19 mortality do not correlate with BCG higher coverage. The implication of this observation for COVID-19 epidemiology is evident.

It is likely that in countries where the young and potentially also the middle-age populations are BCG vaccinated, maximal protection is provided. Universally, the DPM among young people is very low (0.04% for <17 years (*33*)), and their main contribution is expressed in sustaining the chain of infection. Furthermore, the middle-age group (25-64 years) that overlaps with the working-age group is likely to contribute to the spread of the virus is active in a cross-generation interaction (*34*). Thus, higher coverage of young and middle-aged people that leads to a lower death toll in the elderly population is most probably due to a lower rate of infection that is caused by these age groups, leading to an overall lower death toll. The lack of correlation between BCG coverage of >65 years and COVID-19 outcomes is in accord with the expected negligible impact of the elderly population on the transmission within the community. Several reports proposed that BCG vaccinated populations are partially protected against viruses in general, and against the SARS-CoV-2 in particular (*35*). Despite the broad usage of BCG for almost a century, the long-term effect of BCG on the immune system remains enigmatic. Indirect effects of BCG on diabetes, multiple sclerosis, and all-cause mortality of children in developing countries were reported (*36*) and was considered a heterologous effect (*37*). A BCG-dependent reduction in asthma symptom prevalence of children in the UK suggests its impact on the protection of the respiratory system (*37, 38*). We postulate that the positive effects of BCG immunization on COVID-19 outcomes are achieved by an improved systemic immunity of the young population (<24 years). It was also postulated that other types of immunizations are linked to the capacity of the immune system to protect from viral infection. Specifically, the recovery from warts which are caused by human papillomavirus (HPV) was maximal by MMR (measles, mumps, rubella vaccine) vaccination (*39*). We found no correlation between the degree of Measles and Rubella vaccination coverage (within MMR vaccination) and COVID-19 outcomes (**Fig 7**), whereas BCG vaccination coverage resulted in a substantially improved outcome for COVID-19. Recall that all tested countries are signified by quite high coverage for measles (>78% coverage, **Fig. 7b**).

The strong correlation of COVID-19 with the countries which applied recent immunization (for 0-15 years) relative to countries that have only partial or no immunization in recent years is of special interest. The strong statistical significance value for BCG being effective for the recently immunized population (**Fig. 7a**) supports the notion that active immunization contributes to a better COVID-19 outcome (*40*). It was proposed that immunization by BCG acts as a stimulator of the immune system by activating T-helper cells (CD4+) that in turn reports CD8+ cells, NK, and dendritic cells for fighting against viruses. Such heterologous BCG-dependent T-cell activation can contribute to the nonspecific BCG effects by the pretrained immunity system. Many COVID-19 patients are signified by leukopenia and a reduction in CD4+ cells (*41*). The kinetics of an increase in T-lymphocyte was considered as a marker for the recovery in non-severe cases of COVID-19 (*42*). While linking T-lymphocyte, BCG, cell activation and clinical manifestation of COVID-19 needs further validation, it suggests a connection to the short and long-term impact of BCG immunization on the potency of the immune system.

We conclude that the strong negative correlation with BCG administration years, the impact of a recent vaccination, and the validated role of the young population in the spread of COVID-19 are fundamental observations for revisiting global and national BCG immunization policy. While, at present, the WHO does not recommend BCG vaccination for prevention of COVID-19, several clinical trials with BCG are undertaken (*43, 44*).

## MATERIALS AND METHODS

### Data extraction

Information regarding COVID-19 outcomes was extracted daily between January 29th and May 5th, 2020, from the Worldometers website (*33*). Countries were included in the selected cohort if their population is >3M, and they met the criteria of >=3 deaths per 1M population on April 17th. The countries cohort can be seen in Supplementary **Table S1**. Data normalization was performed for each country by accounting for the relevant population size. We included in the analysis of death per million (DPM), cases of hospitalization with serious and critical (SPM) and cases per million (CPM). Demographic measures of countries were extracted from the Worldometers website on April 17th, 2020 (*45*). Information regarding the share of population >65 years and economic development indicators was extracted from the World Bank data (*46*). Prevalence of chronic diseases (e.g. obesity, type 2 diabetes, Tuberculosis), and the death rate from cardiovascular disorders were extracted from Our World In Data website. Supplementary **Table S4** provides the source for each of this country-related information.

Information regarding past and present BCG administration practices in every country were extracted from the BCG world atlas (*45*). Two vaccination status groups were considered. The first group included countries that had either a current or past national mandatory vaccination policy (49 countries). The second group included countries that have only administered BCG vaccinations to specific groups at risk (6 countries). The percentage of the people at risk that were immunized out of the total population size is negligible (*45*). In addition, the estimates for BCG vaccination coverage between years 1980-2018 were extracted from the annual WHO reports (*27*). For additional resources used to establish the years of mandatory BCG administration see Supplementary **Table S3**. The vaccination coverage data for measles and rubella were extracted from WHO reports (*27*).

### Data Analysis

Accounting for the varying time of the diseases across countries, we define a unified aligned key date of a country as the first date when DPM reached for the first time the value 0.5 or higher. The following analysis was conducted across changing dates (e.g. 10-40 days) for the key aligned date as defined. For country-specific groups (binary or categorized, such as BCG administration), we applied the ranked Wilcoxon test as a nonparametric statistical test for comparing any paired groups. For the continuous (i.e. the number of years BCG was administered, the prevalence of TB in the population, median age) data we applied linear regression, reporting the regression fit and the calculated statistical significance (p-value) for the COVID-19 outcomes.

We tested the correlations between outcomes and years of BCG administration using Pearson’s correlation, and reported P-values as well as permutation tests’ p-values (Supplementary **Table S2**.

A correlation between the BCG by age groups was determined by partitioning the population of each country into three groups: (i) 24 years and younger; (ii) 25 to 64 years; (iii) 65 years and older. From the age partition and the BCG coverage within each age group, a value that measures the *percentage of the population in the share of the age group with BCG* is calculated.

### Data and materials availability

All data needed to evaluate the conclusions in the paper are present in the paper and/or the Supplementary Materials. Additional data related to this paper may be requested from the authors. An online tool for displaying the analytical results is available at: https://covi.shinyapps.io/COVID19/. Code and data is available at: https://github.com/nadavrap/COVID19.

## Data Availability

An online tool for displaying the analytical results is available at: https://covi.shinyapps.io/COVID19/. Code and data is available at: https://github.com/nadavrap/COVID19.

https://github.com/nadavrap/COVID19

https://covi.shinyapps.io/COVID19/.

## ACKNOWLEDGEMENTS

We would like to thank the Interdisciplinary Data Science Research (CIRD), the Hebrew University of Jerusalem, for a partial fellowship to D.K.

## SUPPLEMENTARY MATERIALS

Supplementary material for this article is available.

**Table S1.** Statistics of RCV (Rubella) and MCV (Measles) vaccines and COVID-19

**Table S2.** Correlations and p-values of regression of years of BCG administrations and different outcomes

**Table S3.** Additional resources on BCG administration years by country

**Table S4.** Variable resources used across countries and populations

